# Look before diving into pooling of SARS-CoV-2 samples on high throughput analyzers

**DOI:** 10.1101/2020.08.17.20176982

**Authors:** Jason J. LeBlanc, Glenn Patriquin, Janice Pettipas, Michelle Warhuus, Darren Sarty, Colleen Jackson, Charles Heinstein, James MacDonald, David Haldane, Todd F Hatchette

## Abstract

Given the unprecedented demand for SARS-CoV-2 testing during the COVID-19 pandemic, the benefits of specimen pooling have recently been explored. As previous studies were limited to mathematical modeling or testing on low throughput PCR instruments, this study aimed to assess pooling on high throughput analyzers. To assess the impact of pooling, SARS-CoV-2 dilutions were performed at varying pool depths (i.e. 1:2, 1:4, and 1:8) into test-negative nasopharyngeal or oropharynx/anterior nares swabs matrix. Testing was evaluated on the automated Roche Cobas 6800 system, or the Roche MagNApure LC 2.0 or MagNAPure 96 instruments paired with a laboratory-developed test using a 96-well PCR format. The frequency of detection in specimens with low viral loads was evaluated using archived specimens collected throughout the first pandemic wave. The proportion of detectable results per pool depths was used to estimate the potential impact. In addition, workflow at the analytical stage, and pre-and post-stages of testing were also considered. The current study estimated that pool depths of 1:2, 1:4, and 1:8 would have allowed the detection of 98.3%, 96.0%, and 92.6% of positive SARS-CoV-2 results identified in the first wave of the pandemic in Nova Scotia. Overall, this study demonstrated that pooling on high throughput instrumentation can dramatically increase the overall testing capacity to meet increased demands, with little compromising to sensitivity at low pool depths. However, the human resources required at the pre-analytical stage of testing is a particular challenging to achieve.

## Introduction

In response to the 2019 novel coronavirus (COVID-19) pandemic, caused by severe acute respiratory syndrome coronavirus 2 (SARS-CoV-2), there has been an unprecedented demand for laboratory testing. Molecular methods such as real-time reverse transcription polymerase chain reaction (RT-qPCR) have been the primary testing method and a fundamental tool in patient management and public health containment and mitigation strategies.[1–5]

In the initial stages of the pandemic, testing was restricted to symptomatic individuals with a compatible travel history to an area of COVID-19 concern; however, our understanding of SARS-CoV-2 epidemiology improved and the recommendations for testing evolved over time. With global spread and data suggesting the possibility of SARS-CoV-2 transmission through asymptomatic or pre-symptomatic individuals, travel requirements or presence of compatible symptoms were no longer a prerequisite for testing.[6–9] Subsequently, the demand for testing became overwhelming, and laboratories were challenged by insufficient testing supplies.[1,10–13] As COVID-19 cases began to decline, the pressure on laboratories to meet testing demands continued, as testing remained a cornerstone to support the reopening of the economy and easing of public health restrictions.[14,15]

A possible strategy to increase testing capacity and gain laboratory efficiencies, is group testing (i.e. specimen pooling). While many pooling permutations are possible, its simplest application involves combining patient samples prior to testing, and retesting of individual specimens following identification of a positive pool.[16–23] The optimal number of specimens within pools (i.e. pool depth) can be estimated through mathematic modeling, and varies with disease prevalence and assay performance.[16–38] While larger pool depths may achieve higher efficiency, the trade-off is reduced sensitivity and the potential generation of false negative results.[18–23, 31–38] When prevalence is low, typically only a subset of specimens with low viral loads pass undetected.[18–23, 31–38]

Pooling using RT-qPCR has been applied for surveillance of various infectious diseases in both animals and humans.[39–42] For SARS-CoV-2, pooling has been applied in modelling or for relatively small throughput instruments with varying pool depths [16–38], but no studies have described the impact of pooling on automated high throughput analyzers. This study validated pooling of specimens for SARS-CoV-2 testing at different pooling depths on high throughput analyzers, and discusses some practical considerations prior to implementation.

## Methods

### Pooling efficiency modeling

The impact of prevalence and pooling depth on testing capacity was modeled as previously described [17] with online software (https://www.chrisbilder.com/shiny), using an assumption of RT-qPCR sensitivity of 95% and specificity of 99.9%. Testing capacity was calculated for prevalence values spanning 0.1 to 10%, and for pooling depths ranging for 3 to 10. A value above 100% indicates testing capacity is increased.

### Specimen collection and pooling

The collection of specimens for SARS-CoV-2 RT-qPCR testing were performed using a flocked nasopharyngeal (NP) swabs collected in 3.0 ml of universal transport medium (UTM) (Copan Diagnostics Inc., Murrieta, CA), or a Aptima Multitest swab (Hologic Inc., San Diego, CA) for oropharynx/anterior nares (OP/Na) collection in 2.9 ml of specimen transport medium (STM).[10] Each specimen was stored at 4°C until testing, and aliquots were stored at –80°C. For pooling, specimens were diluted in triplicate using a Voyager 8-channel adjustable tip spacing pipette (Integra Biosciences Corp., Hudson, NH) at ratios of 1:2, 1:4, and 1:8, to achieve volumes of 1.5 ml. Dilutions were performed using negative matrix (UTM or STM), consisting of combined SARS-CoV-2 test-negative specimens (n≥48).

### Nucleic acid extraction and RT-qPCR

Swabs material and dilutions were processed with one of three RT-qPCR methods. First, the SARS-CoV-2 assay was used on the automated Cobas 6800 system (Roche Diagnostics, Laval, QC). For NP specimens in UTM, 600 μ1 was processed directly, but for the OP/Na swabs in STM, 200 μ1 was diluted into 1 ml of Cobas omni Specimen Diluent prior to use.[10] Threshold cycles (Ct) values were categorized as positive with dual target positive results (E gene and Orf1a), indeterminate for single target results, or negative in absence of Ct values. The two remaining methods used different extraction methods where a Total Nucleic Acid (TNA) extraction was performed on either a Roche MagNApure LC 2.0 or MagNAPure 96 instrument, and paired with a laboratory-developed test (LDT) [1,10,11]. Briefly, TNA was extracted from 200 μl of specimen, eluted into 50 μl of elution buffer, and 5 μ1 was used as template in a duplex RT-qPCR targeting the SARS-CoV-2 envelope (E) [43] and RNA-dependent RNA polymerase (RdRp). Amplification was performed on an Applied BioSystems 7500 Fast system (Thermo Fisher Scientific, Mississauga, ON). Results were categorized as positive with dual target positive results with Ct values ≥38, indeterminate for single target results between 35 and 38 (unless confirmed by another method with a different genetic target), or negative for Ct values ≥38.

### Estimated impact of pooling on SARS-CoV-2 detection

For each instrument and swab type, a limit of detection (LoD) analysis was performed using 10-fold serial dilutions of previously positive SARS-CoV-2 specimens in UTM or STM, in comparison to viral dilution at varying pool depths (i.e. 1:2, 1:4, and 1:8). Virus concentration was estimated in relation to a standard curve with quantified virus provided by the National Microbiology Laboratory (NML) (Winnipeg, MB).[31] Ct values for specimens dilutions at different pooling depths were compared to undiluted specimens, and used to defined the subsequent analyses.

The frequency of detection in specimens with low viral loads (i.e. Ct values near the defined in the LoD for each pool), was assess retrospectively using archived specimens at −80°C. –80°C. The proportion (%) of detectable results per Ct value category was used to estimate the potential impact of pooling in previous results obtained in Nova Scotia from January 24th, 2020 to June 26th, 2020. Results were categorized by Ct values, instruments, and RT-qPCR targets.

### Considerations for specimen workflow, turnaround times, and human resources

Using direct observation, the average (n = 10) hands-on times tasks required for each instrument was estimated, the theoretical daily maximum specimen throughput was estimated assuming ideal conditions, with no restrictions for human resources, reagents/consumables, or cost. Maximal testing capacity assumed retesting of individuals specimen from positive pools would occur on a separate instrument with equivalent sensitivity [1,17,44–47]. Human resources required for analytical, and pre and post-analytical processing were estimated. Pre-analytical steps accounted for the specimen registration into the hospital laboratory information system (LIS) (i.e. 3–5 min/specimen), labelling, and aliquoting. Analytical steps included specimen organization, pooling, and any instrument pre-processing steps. Post- analytical steps included result interpretation, reporting, and notifications to ordering physicians, infection prevention and control, and public health.

## Results

### Epidemiology and impact of prevalence on testing capacity at various pool depths

In Nova Scotia, the overall daily positivity rates varied from 0.0% to 8.0%, and daily fluctuations were evident (Figure 1A). Using mathematical modelling, it was demonstrated that pooling depths between 3 and 8 were inefficient at a prevalence of ≥ 8%, and pools of 9 or 10 were inefficient at a prevalence of ≥ 6% (Figure 2). However, at low prevalence, testing capacity increased with pool depth.

**Figure 1.**
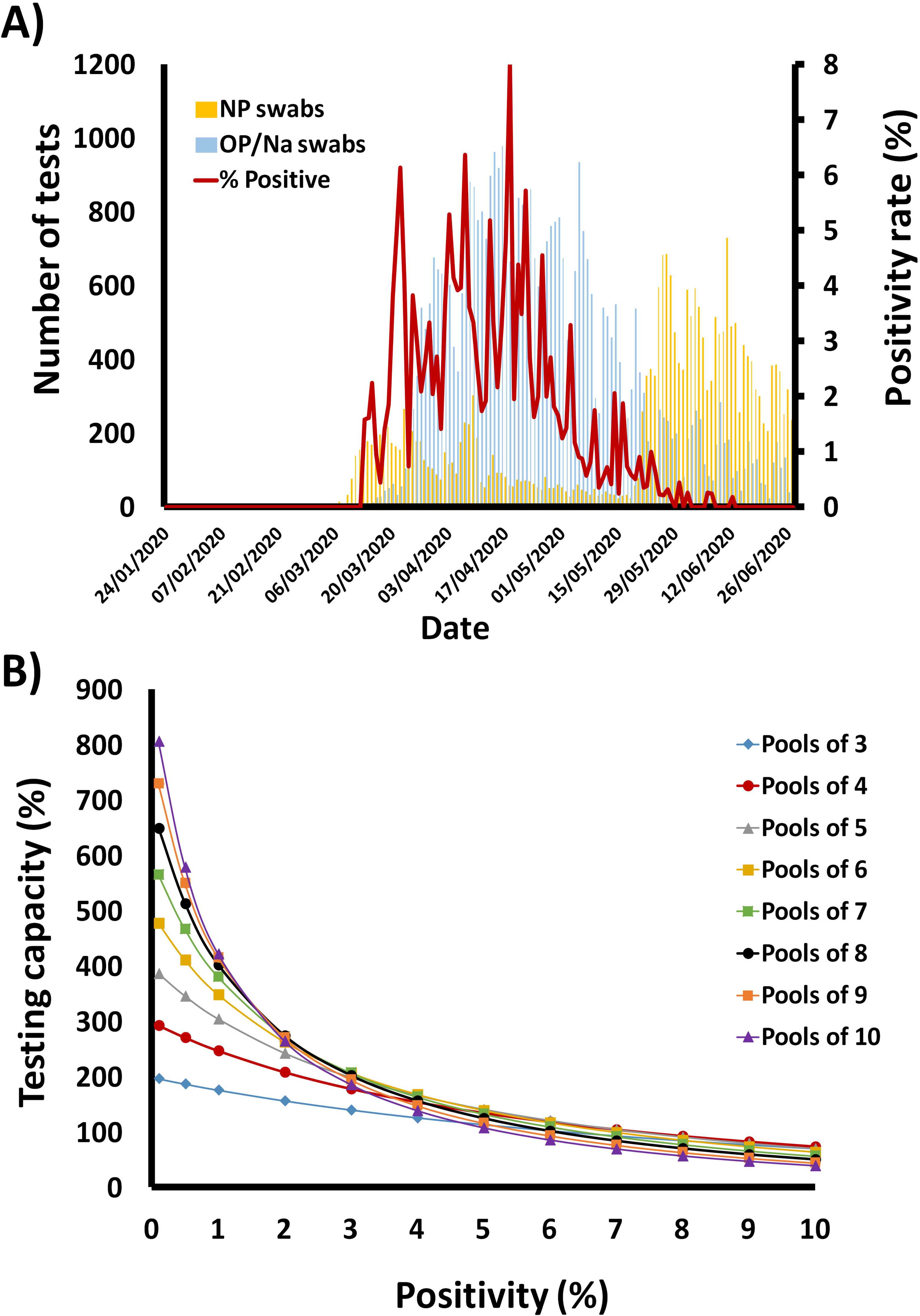
Epidemiology of SARS-CoV-2 in Nova Scotia, and impact of prevalence and pool size on testing capacity. A) The total number of SARS-CoV-2 tests performed (histograms) and positivity rates (in red) are plotted against a time. NP swabs in UTM are depicted in yellow, and OP/Na swab in STM are in blue. B) Testing capacity is plotted against the positivity rate for each pool depth. A value of above 100% was considered an increase in testing capacity, whereas values below 100% were deemed inefficient.

**Figure 2.**
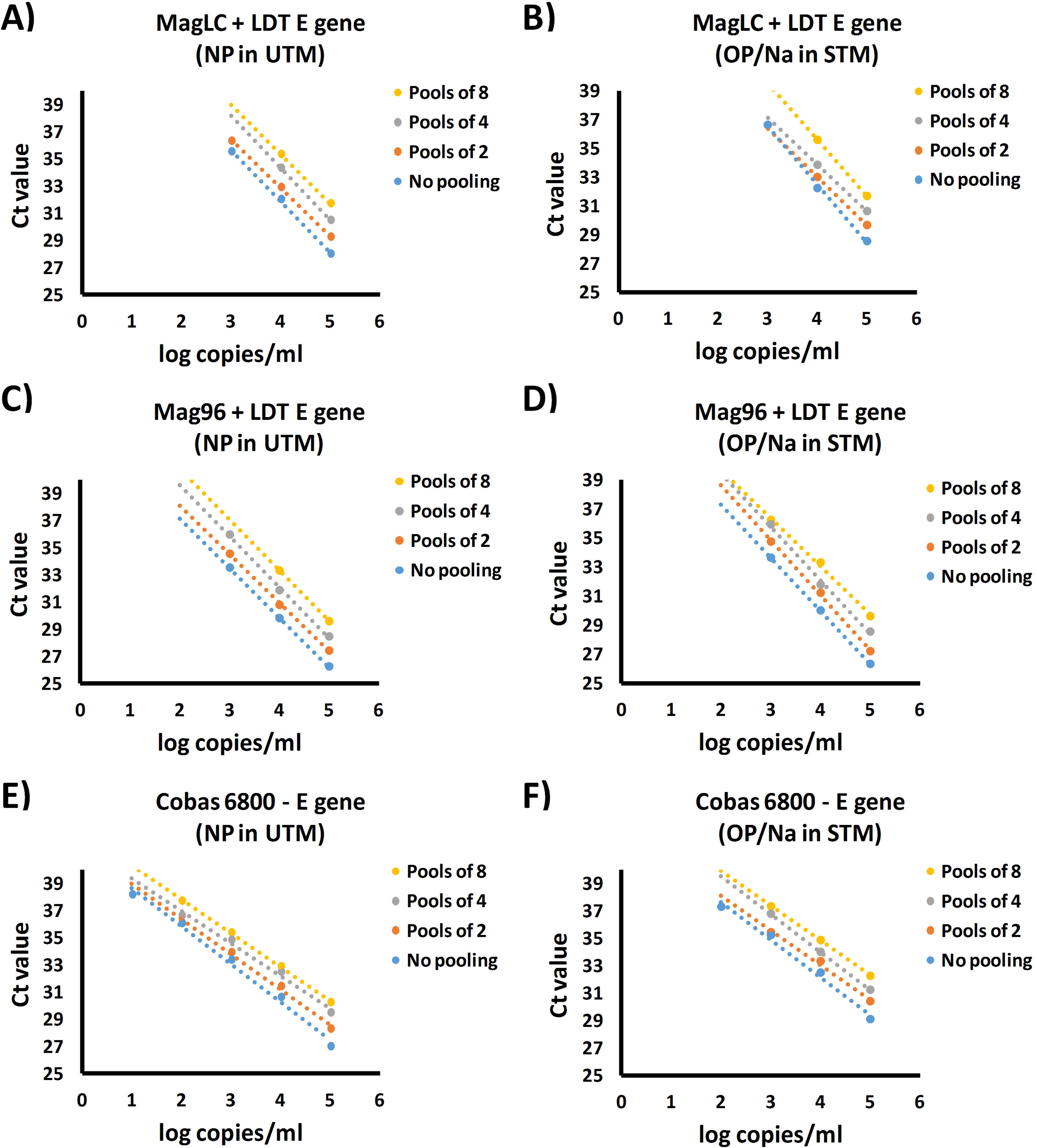
Impact of pool depth on the analytical sensitivity. Ten-fold serial dilutions of SARS-CoV-2 were tested on each instrument and for each swab type, and compared to the same dilutions processed using pooling depths of 1:2, 1:4, and 1:8. Note: The results for the E gene target for each instrument are shown, but similar results were observed for the other RT-qPCR targets: RdRp in the LDT assay, or Orf1a for the cobas 6800 (data not shown).

### Estimated impact of pooling on SARS-CoV-2 detection

Using LoD analyses, the potential impact of pooling at depths of 1:2, 1:4, and 1:8 was assessed for each instrument (Figure 2). Compared to undiluted controls, increasing pool depths progressively reduced the analytical sensitivity, as seen by the decrease in SARS-CoV-2 RNA detection near the LoD, and the incremental increase of approximately 1 to 2 Ct values for each pooling dilution (Figure 2). Similar trends were noted for both NP and OP/Na swabs collections on each instrument (Figure 2). It should be noted that OP/Na specimens required an initial pre-processing step (i.e. 1:6 dilution in manufacturer diluent) on the cobas 6800 instruments and, NP swabs in UTM therefore achieved lower LoDs.[10] Moreover, the cobas 6800 was more sensitive than the LDT paired with the MagNAPure 96and processing on the MagNAPure LC was the least sensitive method. Regardless of the comparative differences in analytical sensitivity, similar trends were noted for the relative reduction in sensitivity with increasing pool depths compared to the undiluted controls.

To further quantify the potential impact of pool depth, the frequency of detection was compared against previously tested specimens with low viral loads (i.e. Ct values ≥ 33 on the cobas 6800 or Ct values ≥ 32 on the LDT). Of 134 archived specimens retested on the cobas 6800, 113 yielded a detectable signal and Ct values were categorized (Figure 3A). Similar approaches were used for the LDT (Figure 3C). Overall, the frequency of SARS-CoV-2 detection with high Ct values decreased as the pooling depth increased (Figure 3). Using these proportions, the number of specimens that would have been missed during the first pandemic wave was estimated (Figure 3B and D). Excluding the 21 indeterminate results that were not reproducible, the proportion of specimens missed for pooling depths of 1:2, 1:4, and 1:8 would be 1.6% (9/570), 5.3% (30/570), and 11.1% (63/570), respectively (Figure 3B). Similarly, if the 17 indeterminate results were excluded for the LDT data, the proportion of specimens missed for pooling depths of 1:2, 1:4, and 1:8 would be 1.8% (12/677), 3.0% (20/677), and 4.4% (30/677), respectively (Figure 3D).

**Figure 3.**
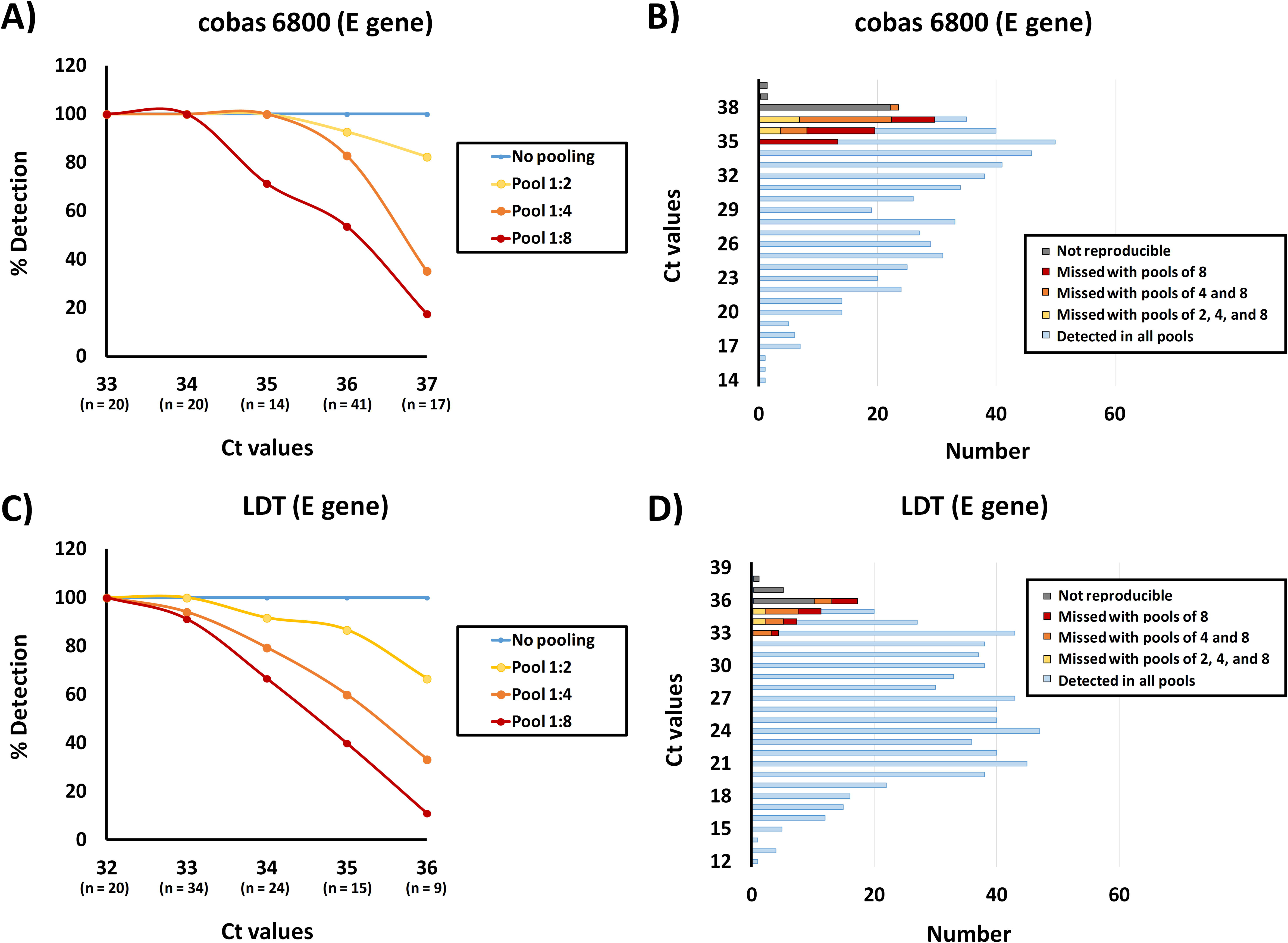
Impact of pooling on the detection of low SARS-Cov-2 viral loads. A) The frequency of detection in specimens with low viral loads was assess using previously tested specimens. B) The proportion (%) of detectable results per Ct value category obtained in A) applied to estimate the potential impact of pooling a depths of 1:2, 1:4, and 1:8. Results were categorized by instruments and RT-qPCR targets, and the E gene target results are depicted. Results for the alternative targets (i.e. Orf1a on the cobas 6800, or the RdRp target on the LDT) showed similar trends as E gene for each instrument (data not shown).

### Considerations for workflow and maximal specimen throughput

The theoretical maximal daily specimen throughput for each instrument was estimated, and increased with pool depth (Figure 4). In terms of specimen processing, the FTEs required in the pre-analytical steps were prominent, and increased with pool depth. These were primarily attributed to specimen registration (Figure 5). The FTE requirements in analytical phase were unchanged by pool depths, and only a small increase would be required in the post-analytical stage given the manual steps required result interpretation, reporting, and communication.

**Figure 4.**
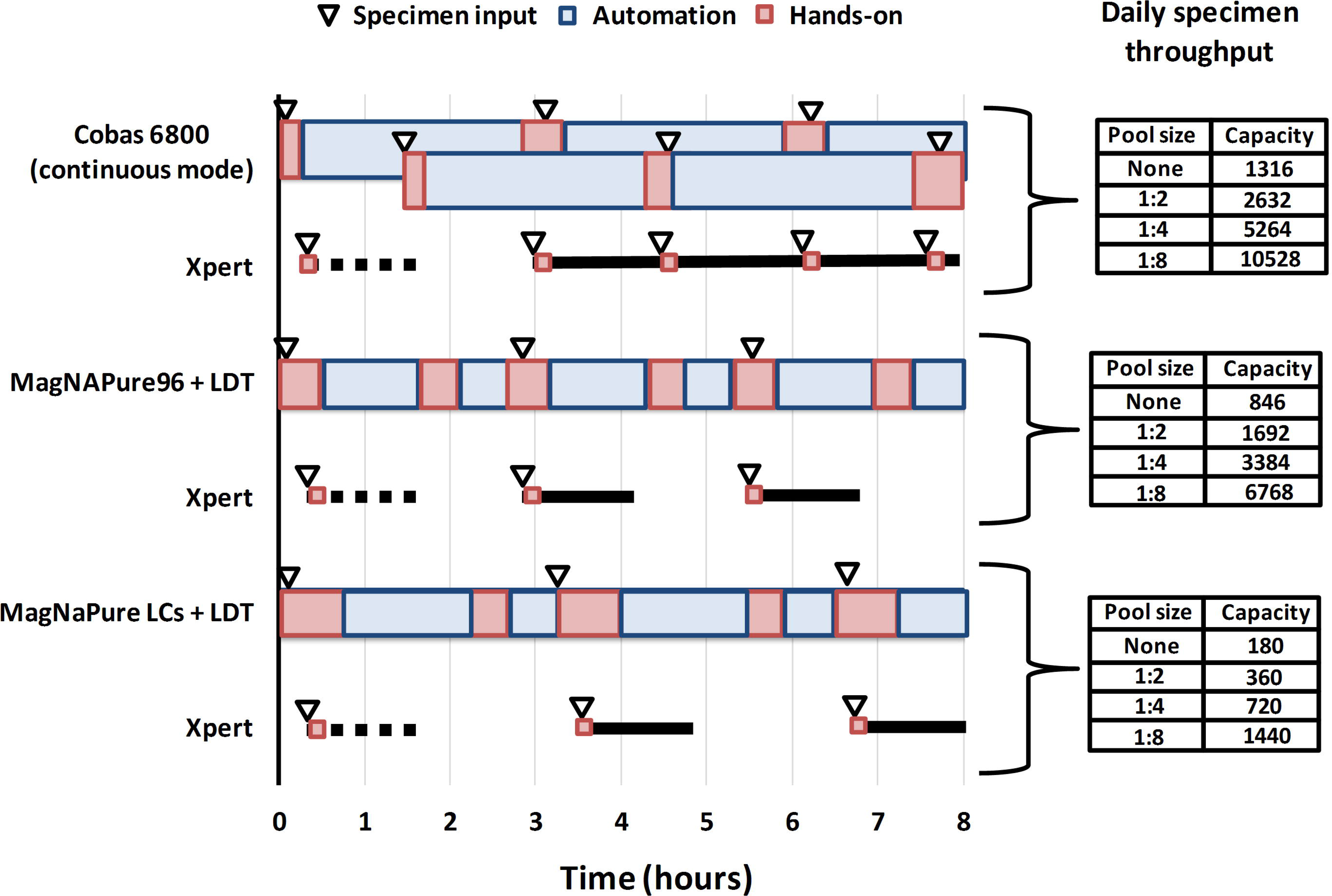
Estimated workflow for maximal specimen throughput on each study instrument based on pool depth. Hands-on time (red), automation (blue), and times required specimens to be loaded onto the instrument (triangles) are illustrated. Ideal workflow assumed resolution of positive pools using a secondary method with equal or greater sensitivity (e.g. Cepheid Xpert Xpress SARS-CoV-2 assay, annotated “Xpert” on the figure.

**Figure 5.**
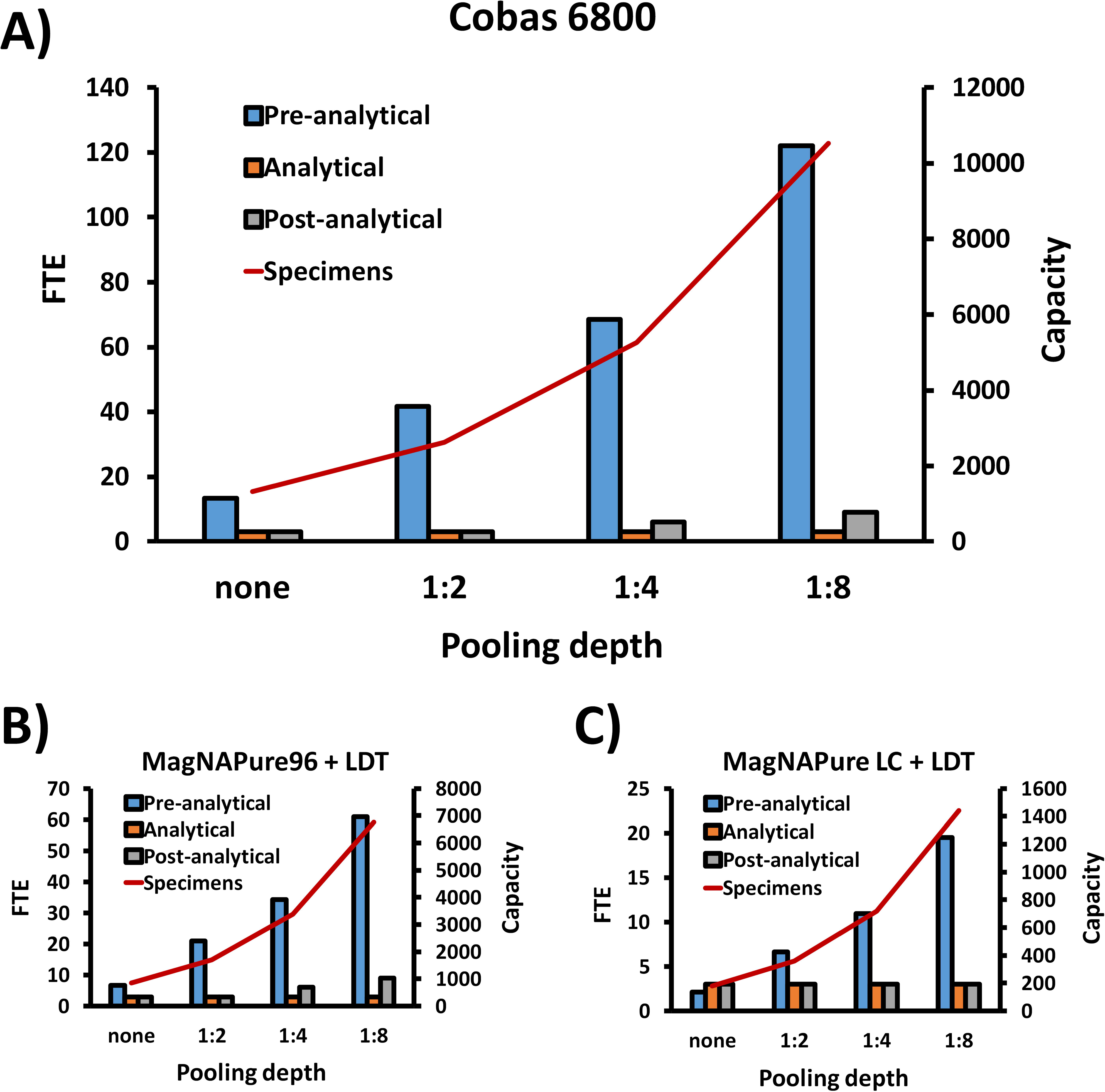
Estimated FTE requirements to achieve maximum instrument capacity. By monitoring the time required for routine testing activities (n = 10), the average time required for each tasks involved at the pre-analytical, analytical, and post-analytical stages of testing were estimated for each instrument, and expressed as the number of full-time employees (FTE) required to support testing at different pool depths.

## Discussion

Given the unprecedented demand for COVID-19 testing, the benefits of pooling have recently been explored for the diagnosis of SARS-CoV-2. To date, pooling for SARS-CoV-2 has been limited to mathematical modeling or testing on low to moderate throughput RT-qPCR instruments.[16–38] In this study, pooling assessed on high throughput analyzers was shown to increase testing capacity with minimal reduction in analytical sensitivity at low pool depths. This study also considered the potential impacts of pooling at the pre- and post-analytical stages. Overall, the theoretical capacity of high throughput instruments could attain over 5,000 tests a day with a conservative pool depths of 1:4, but the human resource required in the pre-analytical stage would be the most significant barrier to implementation.

Since first proposed by Dorfman in 1943 [16], pooling has been well recognized as a strategy to gain efficiency and increase testing capacity.[*] Mathematical models have been developed to help choose the optimal pool depth that would achieve maximum testing capacity, for a defined disease prevalence and instrument performance characteristics.[16–23] Using modeling data for the Cepheid Xpert COVID-19 assay, Becker *et al*. [17] suggested that a pool size of 6 was ideal at a positivity rate of 3%, but at 7.6%, a pool size of 3 would be more efficient. However, in this study, when using modelling to assess the value of pooling at a given prevalence, and establishing a cutoff to decide when to switch between different pooling depths, it was shown that fluctuations in the daily positivity rates needed to be considered. For example, during times of the peak detection of SARS-CoV-2 cases in Nova Scotia, the daily positivity rates fluctuated between 2 and 8%. At these rates, and using a conservative pooling depth of 1:4, testing capacity would range between being efficient (at 208%) and being inefficient (at 92.3%). However, rates as high as 8% were infrequent, so a pool depth of 1:4 would have been efficient throughout the entire pandemic wave with the exception of a single day that reach a positivity of 8%. Even a pool depth of 1:8 would have been efficient for all but 3 days, when prevalence exceeded 6%. Therefore, a significant amount of resources could have been saved by pooling in Nova Scotia.

Whether used to meet increased testing demands or as resource sparing strategies, the benefits of each pool depths must be balanced against the potential concomitant loss in sensitivity. Typically, in times of high disease prevalence, pool depth is kept as low as possible to achieve desired testing capacity. In times where prevalence is low (i.e. ≤ 2%), higher pool depths could be tolerated to conserve reagents in preparation for subsequent waves of COVID-19, or as a mechanism to increase testing capacity to support broader surveillance strategies.[8,15,28–34] This study, like others, showed that higher pool depths increase the risk of false negative results, as the inherent loss in sensitivity fails to identify a proportion of specimens with low viral loads.[17,18,21,23,31,33,36] Typical pool depths used for SARS-CoV-2 RT-qPCR testing range from 3 to 12 [17,31,37,38], yet higher pool depths have been attempted [18,21,23,26,34]. In an extreme example of high pooling depth, Hossain *et al*. [48] released an unpublished document that describes pooling of up to 19,200 RNA samples for simultaneous detection of SARS-CoV-2 using next-generation sequencing (NGS). While NGS technology could be a powerful tool for disease surveillance, this protocol was not validated for clinical testing, and the authors did not consider the significant loss of sensitivity that would likely occur from such a substantial level of pooling.

The extent to which specimens are pooled is predicated on the level of risk that is deemed acceptable. For example, the Canadian Blood Services (http://www.blood.ca) commonly used a pool depths of 1:6 for bloodborne pathogen screening using RT-qPCR, to ensure adequate sensitivity even if the overall disease prevalence is low. Overall, the current study estimated that pool depths of 1:2, 1:4, and 1:8 would have allowed the detection of 98.3%, 96.0%, and 92.6% of positive SARS-CoV-2 results identified in the first wave of the pandemic in Nova Scotia. Without a prospective comparison, it is difficult to ascertain the true impact of the SARS-CoV-2 cases with low viral loads that would potentially have been missed with pooling. Specimen pooling should have a minimal impact on clinical sensitivity for detection of new symptomatic cases, as high viral loads are generally present during this stage of illness.[7,49–56] Similarly, missing SARS-CoV-2 in individuals with low viral loads in the recovery stage might impact the epidemiologic case counts, which is important for public health contact tracing, but would have little value for patient management as these individuals are likely no longer infectious.[18,36,49–52]. Conversely, undetected low viral loads may represent asymptomatic individuals or those in early or late stages of infection.[49–56] To mitigate the risk of missing early or pre-symptomatic infections with pooling, individuals should be encouraged to self-monitor for symptoms, self-isolate, and undergo repeat testing at defined time points.[5,8,14,15,17,28,29] On a population level, a combination of pooling, mass testing, and repeat testing could help cases that would otherwise not have been captured with routine individual testing.[8,14,15,28,32–34] In other words, the decreased sensitivity that is inherent to pooling could theoretically be offset by the reciprocal increased testing capacity, if mass testing and repeated testing over time would improve case finding.

From an analytical standpoint, testing capacity increases with pooling depth if prevalence is low, and the risk of generating false negative results with pooling is minimized by using the most sensitive method available. In this study, both the analytical sensitivity and daily specimen throughput was shown to be highest on the Cobas 6800, and specimen throughput increased with pool depth. The impact of high prevalence on testing capacity could be reduced by retesting positives pools on a secondary instrument, rather than resolution of pools on the subsequent run on the same instrument; however, the secondary assay should have equal or greater sensitivity. The Cepheid Xpert Xpress SARS-CoV-2 is a rapid molecular assay has been shown to have comparable performance to the Cobas 6800 [10,31,44–47], but was not assessed in parallel in this study due to the limited availability of tests. In times of low prevalence and reduced test numbers, positive pool resolution could easily be accommodated on the same instrument used for initial pool testing; however, lower throughput analysers could be considered for smaller test volumes, recognizing the relative decrease in sensitivity of these methods would have less of an impact during times of low disease prevalence. Whether in time of high or low prevalence, testing should not be based on maximal instrument capacity, but on established turnaround time goals, to avoid delays in specimen result reporting. Overall, pooling on high throughput analyzers could be undertaken with only minor changes to the analytical workflow.

No previous study has described the potential impacts of pooling at the pre- and post-analytical stages of testing. First, there are a number of factors that influence pre-analytical steps that are outside the scope of this study (e.g. specimen type, quality and timing of collection, and transport conditions). Once specimens arrive to the laboratory, routine activities in the pre-analytical stage include registration, labelling, aliquoting, and any pre-processing steps required prior to testing. While automation (i.e. robotics) could be used to enhance specimen traceability, reduce the potential for contamination, and help with specimen organization [20,23], the biggest contributor to workload in the pre-analytical stage in our laboratory is specimen registration into the laboratory information systems (LIS). The LIS eventually communicates the test results to the ordering physician, and other healthcare providers (e.g. infection prevention and control, and public health). Specimen registration is crucial to all laboratory testing. Meeting the FTE requirements that accommodate higher testing capacity is one of the biggest barriers to pooling on high throughput analyzers in our laboratory. In contrast, at the post-analytical stage, FTE requirements required to increase testing capacity would be subtle, and largely dependent on the time required for result interpretation, reporting, and communications. These processes could potentially be streamlined with automation if the instruments are interfaced to the LIS. Future studies will explore the incremental benefits of automation at the pre- and post- analytical steps, paired with specimen pooling on high throughput analyzers.

This is the first study reporting the performance of pooling on a high throughput analyzers, with considerations for workflow at the analytical stage, and pre-and post-stages of testing. Assuming reagent availability, the most significant barrier to implementation of pooling in our laboratory is not the instrumentation, but the number of FTEs required to support specimen collection and registration. Careful consideration should be given to all aspects of testing prior to implementation of pooling.

## Data Availability

All data is available in the content of the manuscript

## ACKNOWLEDGEMENTS

This research received no specific grant from any funding agency in the public, commercial, or non-for-profit sectors. The authors have no conflicts to declare. The authors would like to thank the members of the Division of Microbiology, Department of Pathology and Laboratory Medicine, Nova Scotia Health, who were instrumental for sample processing and laboratory testing. The authors are also indebted to the members of the Nova Scotia Department of Health and Wellness, who facilitated data collection and epidemiological analyses during the pandemic.

